# Simulation of COVID-19 Epidemic from Potential Viral Loads in Saudi Arabian Wastewater Treatment Plants

**DOI:** 10.1101/2023.09.30.23296175

**Authors:** Mutum Zico Meetei, Ahmed H. Msmali, Abdullah Ali H. Ahmadini, Shokrya Alshqaq, Hassien M Alna-shiri

## Abstract

SARS-CoV-2 is a contagious respiratory virus that has been discovered in sewage, human waste, and wastewater treatment facilities. Wastewater surveillance has been considered one of the lowest-cost means of testing for tracking the COVID-19 outbreak in communities. This paper highlights the dynamics of the virus’s infection, persistence, and occurrence in wastewater treatment plants. Our aim is to develop and implement a mathematical model to infer the epidemic dynamics from the possible density of SARS-CoV-2 viral load in wastewater. We present a log-normal model and fractional order of susceptible-exposed-infected-recovery (SEIR) epidemic model for predicting the spread of the COVID-19 disease from the wastewater data. We study the dynamic properties of the fractional order SEIR model with respect to the fractional ordered values. The model is used to comprehend how the coronavirus spreads through wastewater treatment plants in Saudi Arabia. Our modeling approach can help with wastewater surveillance for early prediction and cost-effective monitoring of the epidemic outbreak in a situation of low testing capacity.

## 1. Introduction

Wastewater-based epidemiology (WBE) is a prominent early pandemic detection and warning approach. Water virus contamination has become a problem of concern since the COVID-19 pandemic broke out. Workers at wastewater treatment plants are susceptible to contracting the coronavirus from the wastewater [1]. Mass testing programs are implemented as a containment measure when the pandemic is still in its early phases. Although the mass clinical testing program yields useful information on public health, maintaining mass testing at the community level necessitates a pricey and sturdy infrastructure with a steady supply of testing supplies and personnel.

Since the COVID-19 virus can be found in the feces of infected people, including those who are asymptomatic, three days after infection [2], public health organizations have recently considered WBE as a powerful tool to predict the infection. SARS-CoV-2 virus load in wastewater is highly correlated with changes in COVID-19 prevalence in nearby communities [3]. Over 72 countries and 288 universities have implemented WBE for COVID-19 since the pandemic’s start [4]. In Arab nations, little research has been done on wastewater approaches. In United Arab Emirates (UAE) and Saudi Arabia, the SARSCov-2 has been identified and found in wastewater [13, 14]. Wastewater-based epidemiology may be useful for COVID-19 community surveillance, according to the discovery of SARS-Cov-2 in wastewater [15]. In Saudi Arabia, the wastewater approach for assessing coronavirus spread is rarely employed. Several research [16, 17, 18] used wastewater surveillance reports to track COVID-19 infection in the community.

Prior to changes in local COVID-19 mass testing, all SARS-CoV-2 wastewater surveillance reports tracked the same changes [5-7]. Epidemiology based on wastewater can also be used to forecast hospitalizations and ICU admissions and evaluate the success of public health initiatives [8,9]. Without intensive clinical surveillance, WBE can estimate coronavirus and other important disease transmission parameters in the community in addition to monitoring trends. In the absence of clinical surveillance, the mathematical model and the data on viral shedding in wastewater can be an effective tool for COVID-19 prediction [39].

Many studies have tried to use multiple regression models, mathematical models, and artificial intelligence to estimate COVID-19 prevalence or predict the outbreak through WBE [10-12]. Fractional derivatives are effectively used to explain the natural behavior of epidemics. The two most well-known fractional operators are Caputo and Riemann-Lioville. The Caputo derivatives have the advantage of enabling natural modeling over the fractional derivatives because a constant’s derivative is zero. As a result, utilizing the Caputo fractional differential equations, local initial and boundary conditions can be incorporated into the model derivation [19]. In the fields of science, engineering, and epidemiology in particular, the fractional system is important in studying memory effects and inheritance, which cannot be studied using integer-order models [37]. In this paper, we look at various models that can be used to better comprehend the dynamic behavior of COVID-19 infection and its prevalence in wastewater. A methodology to follow in the process of identifying coronavirus affected regions by using log-normal distribution, and fractional ordered susceptible-exposed-infected-recovered (SEIR) epidemiology models. The simulated results are compared with the real data to validate the models’ accuracy.

The paper is organized as follows: Section 2 provides brief information on wastewater treatment and monitoring. Section 3 presents an overview of the preliminary. Section 4 introduces the mathematical models for the transmission of COVID-19 through wastewater. The findings of this study’s use of the fractional order SEIR model for predicting virus transmission in Saudi Arabia are presented in Section 5. Finally, the conclusion is stated in Section 6.

## 2. Wastewater treatment plant (WWTP) and Surveillance

In a country with a hot climate and scarce natural water supplies, the demand for water has surged for a number of reasons. Despite having lakes and rivers, Saudi Arabia ranks third globally in terms of water consumption per person [25]. The major Saudi Arabian wastewater treatment plants (WWTPs) treatment technique is shown in Table 1.

**Table 1.** Saudi Arabia’s WWTPs: their treatment, operation, and purpose.

| Province | WWTPs | Process | Level Types | Uses |
| --- | --- | --- | --- | --- |
| Riyadh | Manfouha | Activated Sludge (AS) | Tertiary (T) | Irrigation |
|  | Hayer | AS | T | Groundwater recharge and Irrigation |
|  | Refinery | Filtration | T | Irrigation |
| Jeddah | Alkhumrah | AS | T | Irrigation |
| Madinah | Al Madinah | Filtration | T | Irrigation |
| Makkah | Makkah | AS/Filtration | T | Irrigation and industrial |
| Taif | Taif | AS | T | Landscape Irrigation |
| Dammam | Qatif | Oxidation ditch | T | Landscape Irrigation |
| Jazan | Jazan city | AS | T | Irrigation |

The understanding of wastewater treatment techniques has improved through time, favoring modern techniques over more conventional ones. These innovative methods make use of mathematical modeling. The biochemical and physical procedures involved in the technical purification of wastewater are described in the models for wastewater treatment plants. A simple mechanism of the wastewater treatment process is shown in Figure 1.

**Figure 1:**
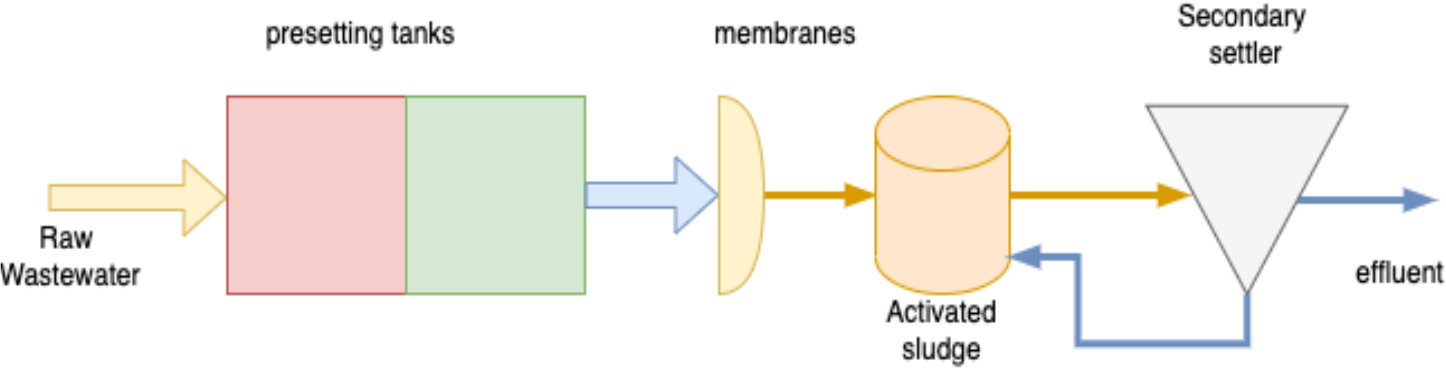
Mechanism for the process in wastewater treatment plants.

The wastewater treatment process consists mainly of presetting tanks, membranes, activated sludge, and secondary settler. The activated sludge model No.1 (ASM1) is the most widely used mathematical model and its updated version has a more advanced description of biological and chemical processes. It is one of the most significant treatment processes, being utilized in around 90% of municipal wastewater treatment processes.

In the beginning stages of the COVID-19 pandemic as well as in the latter stages after it has ended, wastewater surveillance may be a crucial tool [29]. Wastewater surveillance could offer early warning signs and encourage a quick response for locating infected people. The presence of SARS-Cov-2 in WWTP is used as a community COVID-19 infection early warning method. It has been commonly used as a low-cost tool for community-level COVID-19 pandemic monitoring and aids in social and public health decision-making [28]. Figure 2 illustrates a novel approach to controlling and eradicating the COVID-19 outbreak by using wastewater surveillance to achieve accurate epidemic control during a significant outbreak.

**Figure 2:**
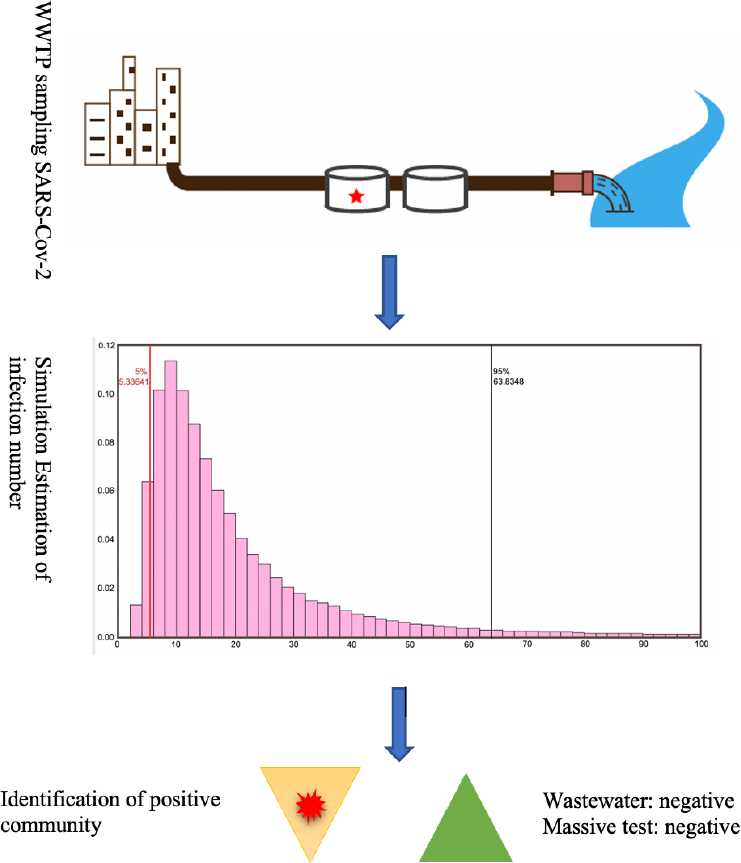
WWTP and community wastewater surveillance as a COVID-19 control plan.

Despite the fact that the detection of clinical samples continues to be the norm for monitoring and tracking, the availability of such data is constrained by bias and the difficulty to track asymptomatic individuals [27]. Quantitative wastewater surveillance might determine the number of infected individuals and provide timely feedback on the effectiveness of the management measures during the development stage. Together, WWTP and community wastewater surveillance could identify any untested omitted infected cases.

## 3. Definitions

In this section, we described some of the foundational concepts of fractional calculus, and log-normal distributions are defined here.

### Definition 3.1

([20]). *For an integrable function G, the Riemann-Lioville fractional-order integral operator can be written as follows:*

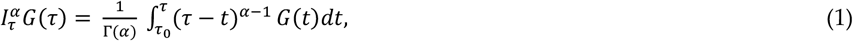

*where τ ≥ τ*_0_, 0 < α < 1, *and Г*(.) *is the gamma function*.

### Definition 3.2

([20]). *For a function G, the Caputo fractional-order derivative operator can be written as follows:*

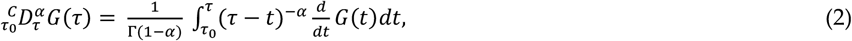

### Definition 3.3

([21]). *Fractional forward Euler method* 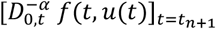 *is approximatedby the left fractional rectangular formula as follows:*

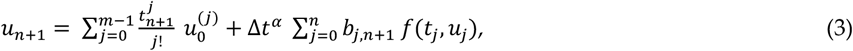

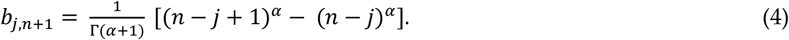

The Definition 3.1 and 3.2 satisfy the following properties:

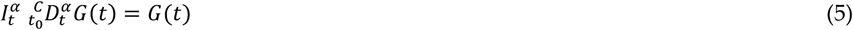

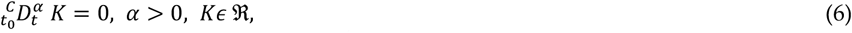

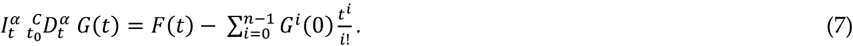

### Definition 3.4

*Log-normal is defined as a continuous probability distribution of a random variable whose logarithm is normally distributed. Suppose Z is a standard normal variable, and mean μ* > 0 *and standard deviation* σ> 0 *be two real number, the log-normal is defined as X*= *e*^5/67^.

To produce a distribution with desired mean *μ*_8_ and variance 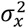 we can be defined as follows:

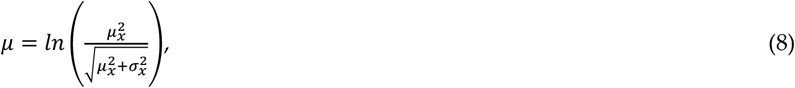

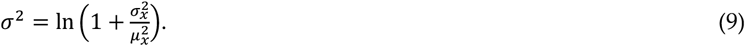

## 4. Material and Methods

We adopted the log-normal distribution and fractional-ordered SEIR model to predict the spread of SARS-CoV-2 through wastewater. The Ministry of Health (MOH) of Saudi Arabia’s website (https://covid19.moh.gov.sa/) provided the theoretical assumptions and data used in this study. We consider the statistical data of coronavirus observed in Saudi Arabia from April 01, 2020 to August 31, 2020 during this time

### 4.1 Normal Distribution

The normal distribution and its characteristics are crucial to the analytical examination of the coronavirus traces found in untreated wastewater from wastewater treatment plants. The data of SARS-CoV-2 traces collected from *N* wastewater treatment plants is deemed to be normally distributed. The equation representing the rate of change in viral load over time *τ* at untreated wastewater plant is given as follows:

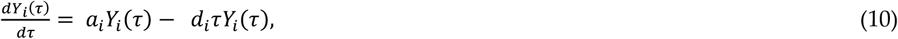

where *d*_*i*_ and *a*_*i*_, respectively, indicate the change in deviation and average deviation of the date based on the daily viral load and concentration in untreated wastewater from *i* WWTP. Let *Y*_*i*_ (*τ*) be the coronavirus trance viral load found in untreated wastewater of *i* wastewater treatment plant, over time interval *τ*_0_ ≤ *τ* ≤ *τ*_*ma*x_. Let σbe the associated standard deviation and *μ* be the average viral load detected in untreated wastewater from WWTP. Equation (10) can be expressed as follows;

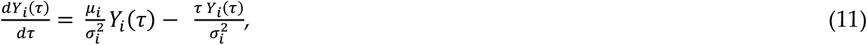

and the exact solution is

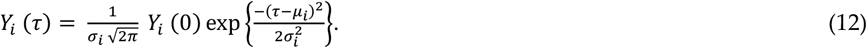

To predict the potential virus load in untreated wastewater facilities, this set of solution (12) can be displayed.

### 4.2 Log Normal Model

Many natural occurrences can be modeled using the log-normal distribution, a crucial statistical distribution. In the log-normal paradigm, modest percentage changes represent numerical growth processes in log scales. The distribution will be more closely resemble normal if these modifications do not have a major impact. We used log-normal distribution to illustrate how the COVID-19 virus spread through wastewater treatment plants. The normal distribution *Z* = ln(*Y*) will then display the dynamic of the COVID-19 spread if the random change *Y* follows a log-normal distribution. For instance, the log-normal distribution will be a characteristic of *Y* = exp(*Z*) if *Z* = exp(*Y*) exhibits the dynamics of a virus spreading. When assuming that the data gathered from the wastewater treatment plants will follow a log-normal distribution, the following mathematical equation is presented:

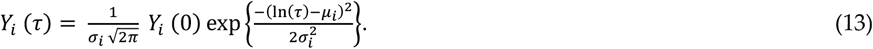

Equation (13) and the gathered data can be compared to find the parameters *μ*_*i*_ and 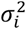as follows:

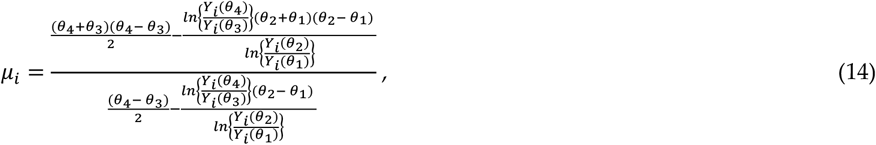

where {*θ*_1_, *θ*_2_, *θ*_3_, *θ*_4_} are the total number of days in the period of time *τ*_0_ ≤ *τ* ≤ *τ*_*ma*x_. We considered the collection of viral loads from untreated wastewater treatment plants for these consecutive days.

The simulation of the log-normal equation (13) for various viral loads in WWTP is shown in Figure 3. It is conceivable to find high SARS-CoV-2 traces because of the large number of affected people, especially when the collection from untreated wastewater facilities was carried out in five major Saudi cities.

**Figure 3.**
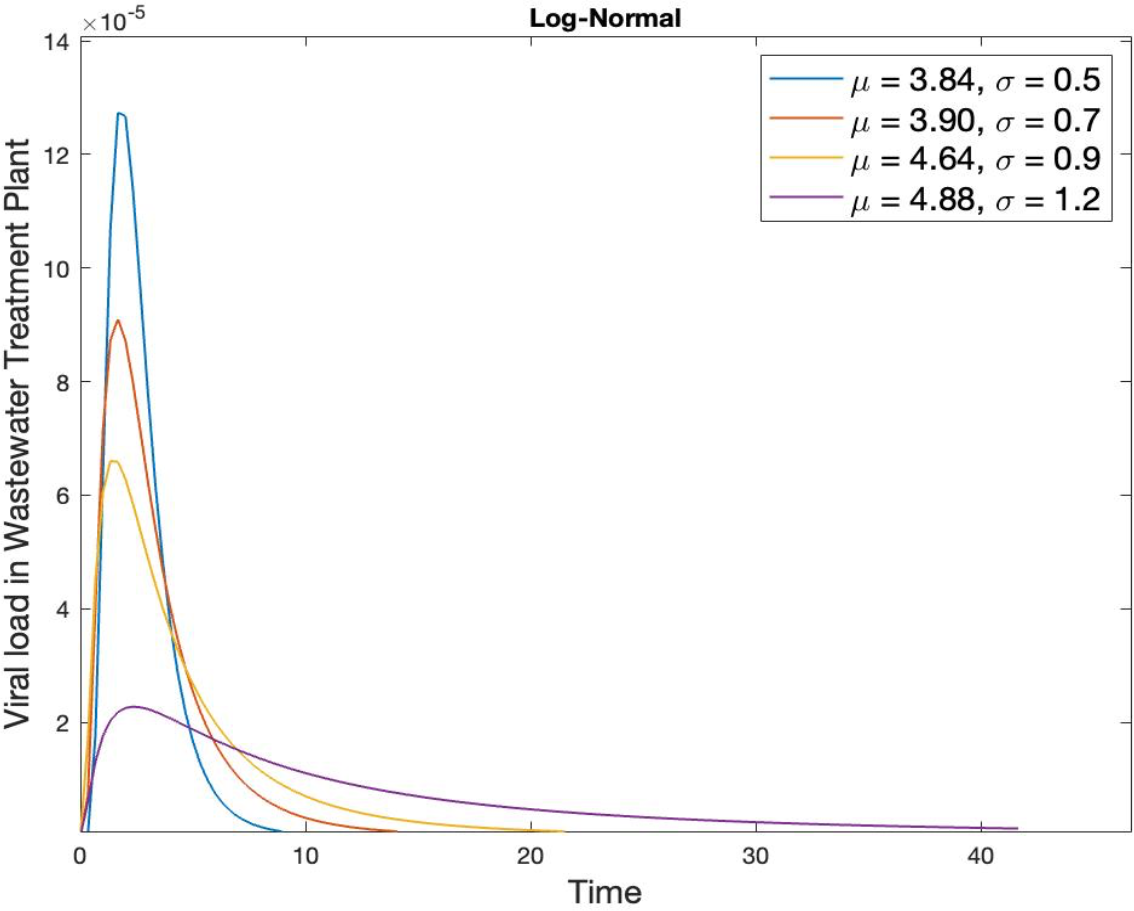
Potential density of SARS-CoV-2 load using Log-Normal model in the WWTP.

In order to ensure that adequate data collection was done, we considered various average (mean) and standard deviation (SD) distributions. In the three untreated sewage facilities in Riyadh, Saudi Arabia, during the months of June, July, and August 2020, the mean viral load was around 1, with the proportion to standard deviation being 0.07, according to the authors of [17]. Plots were made of the data obtained from the major cities’ wastewater treatment facilities through time. Assume a situation where each wastewater treatment facility’s data from the relevant Saudi Arabian major cities has a daily mean (*μ*_1_ = 3.84, *μ*_2_ = 3.90, *μ*_3_ = 4.64, *μ*_4_ = 4.88) and SD (*σ*_1_ = 0.5, *σ*_2_ = 0.7, *σ*_3_ = 0.9, *σ*_4_ = 1.2) that may be represented by a normal distribution.

### 4.3 Susceptible Exposed Infectious Recovered (SEIR) model

To simulate the dynamic behavior of the SARS-CoV-2 virus spreading process through wastewater, the well-known SEIR model was used. In epidemiology, the compartmental pandemic model SEIR is widely used to characterize the COVID-19 outbreak. The concept suggests that the dynamics of virus transmission are determined by the number of populations that are susceptible and infected. When a person has been infected but is not exhibiting symptoms, that time is referred to as the incubation phase. It makes sense to represent the pandemic using a different compartment representing exposed humans without virus spreaders because the coronavirus sickness has a protracted incubation period [22]. In the SEIR model, it is assumed that the population **s** size is fixed and that there are no important dynamics during the long duration. According to their infectious condition, people are divided into four groups or compartments in this paradigm. The detail of the model’s compartments is listed as follows:

- People who are sensitive to the illness but not affected with the disease are referred to be susceptible (*S*).
- People with the virus who are already infected with the virus but are unable to spread are referred to be exposed (*E*).
- The number of infected individuals, the number of infections, and the virus’s capacity to transmit to vulnerable individuals are referred to be infected (*I*).
- Recovered (*R*), refers to the number of individuals who have recovered from infection and
- total population (*N*) = *S*(*τ*) + *E*(*τ*) + *I*(*τ*) + *R*(*τ*).

The following is the simple SEIR compartmental model for COVID-19.

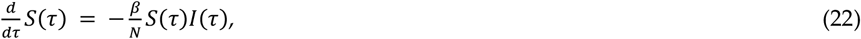

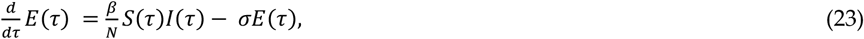

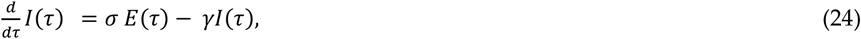

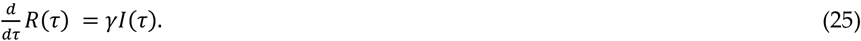

Any numerical method can be used to solve the aforementioned set of equations (22-25), and the behavior of SARS-Cov-2 can be predicted via simulation. The aforementioned system of equations can be expressed when discrete time Δ*τ* is involved, shown as follows:

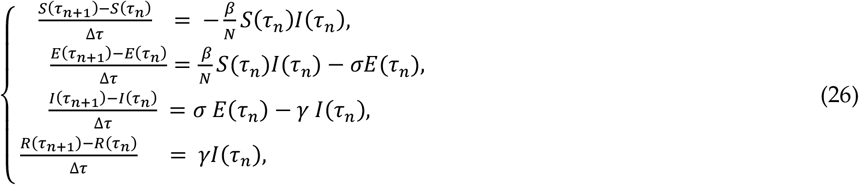

where the description of the parameters can be outlined in Table 1, stated below.

**Table 1.** Definition of the parameters and their values.

| Parameters | Definitions | Values/Dimension | Method and Reference |
| --- | --- | --- | --- |
| $N$ | Total population | 34,218,169 /Individual | [23] |
| $\beta$ | Probability of infection | $9.5 * 10^{-2}$ /Day | Data fitted [23] |
| $\sigma$ | Probability of incubation | $1.7836 * 10^{-5}$ /Day | Data fitted [23] |
| $\gamma$ | Average rate of people recovered over time $\tau$ | 0.96 /Day | Data fitted [23] |

The simulated value of model parameters shown in Table 1 were fitted with values from the Ministry of Health’s official website [23]. We predicated a five-day incubation period as the norm.

#### 4.3.1 Models via Wastewater and COVID-19

The ability to assess the overall viral load in a wastewater treatment plant and keep tracks on the COVID-19 infection in community has been made possible by wastewater base surveillance. However, there are still limitations on measuring and forecasting viral transmission in a community. A small number of recent studies have attempted to use SEIR models to bridge the gap between viral shedding measurements and reported infection cases. For instance, in [30], the authors used SEIR-V model to determine COVID-19 prevalence and infection using wastewater surveillance data. The model proved successful in forecasting the true number of cases 6 to 16 days earlier and higher than the reported number of cases. In [31], the authors used a copula-based time series model to predict COVID-19 infection based on wastewater viral load and clinical reports. The data on the viral load in wastewater were integrated into a stochastic SIR model to investigate and forecast the evolution of the COVID-19 epidemics at the municipal level [32]. To estimate the pandemic for the first wave, the authors in [33] used a two-strain virus competition SIR model fitted using wastewater data and integrating time delay. To forecast the unknowable epidemic data, [34] created a wastewater-based compartmental epidemic model with two-phase vaccination dynamics and immune evasion. A stochastic SEIR model was applied to bridge the gap between the number of infected cases and viral concentration in wastewater [35]. The authors of [36] developed a stochastic SIR model and solved it using the Monte Carlo method to forecast the behavior of the pandemic including viral load data and a variable infection rate to account for the effect of mitigation measures. In order to improve the ability to predict new weekly COVID-19 case numbers, a time-series based machine learning (TSML) strategy was used to extract deeper understanding and insights from WBE data in the presence of other relevant temporal variables, such as minimum ambient temperature and water temperature [36].

#### 4.3.2 Fixed Points and Stability

The fixed points represent disease free conditions. When creating a disease model, it’s essential to determine the endemic equilibria (fixed points) in order to rule out the possibility that one or more equilibria exist [38]. At the beginning of the epidemics in Saudi Arabia in March 2020, the fixed points are

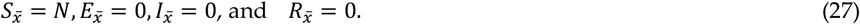

We consider the fixed points of the system (26) as follows:

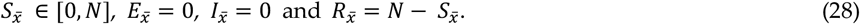

We can find the eigenvalues of the Jacobian matrix after linearizing the system (26) of the fixed point. These eigenvalues can determine the nature of the fixed points at *N* = *N*_0_. The Jacobian matrix at the fixed points is given as follows:

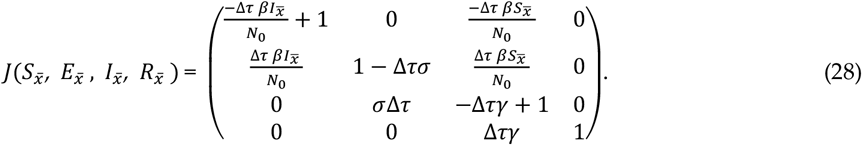

At the fixed points 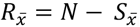, we have

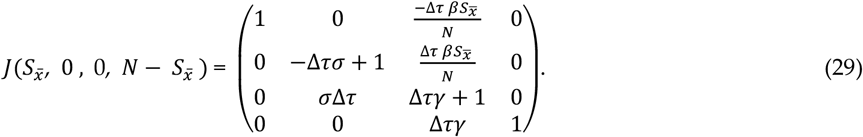

The eigenvalues of the above matrix (29) are given as follows:

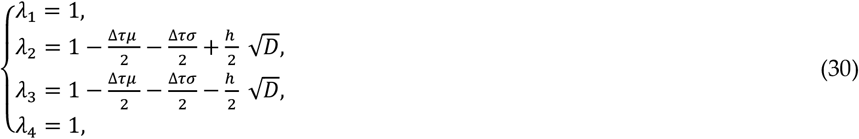

when 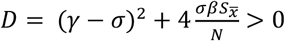 We can show that the fixed points are stable, when|*λ*_2_| < 1 and |*λ*_3_| < 1 ; unstable when |*λ*_2_| > 1or |*λ*_3_| > 1 for *λ*_1_ = *λ*_4_ = 1.

### 4.4 Fractional-Order Susceptible Exposed Infectious Recovered (SEIR) model

It is well known that the adaptation of fractional-order derivative into epidemic model equations can lead to improvement of model parameters in several process. The fractional-order generalization of the SEIR model can significantly improve the model performances. The fractional SEIR model in the sense of Caputo fractional ordered derivative is given as follow:

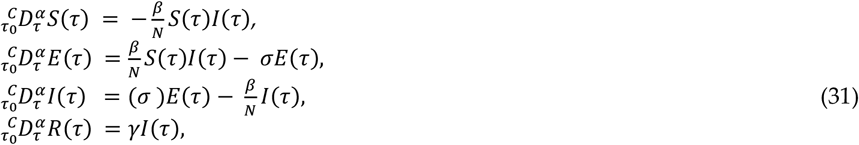

with the initial conditions:

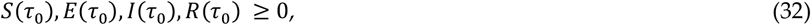

where 0 < α < 1 and 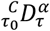is the Caputo fractional-ordered derivative operator.

### 4.4. Existence and Uniqueness

We use fixed-point theory and *Picard-Lindelof* method to derive the existence and uniqueness of the solution of the system (31). The system (31) can be rewritten as follows:

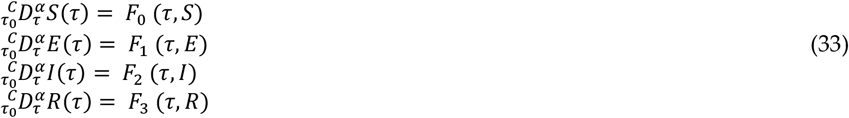

where the functions *F*_4_, 0 ≤ *i* ≤ 3 are defined as follows:

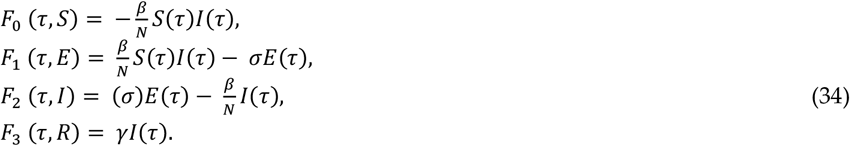

From the initial conditions (32), and applying the fractional-order integral operator (1), the system (31) would transformed as follows:

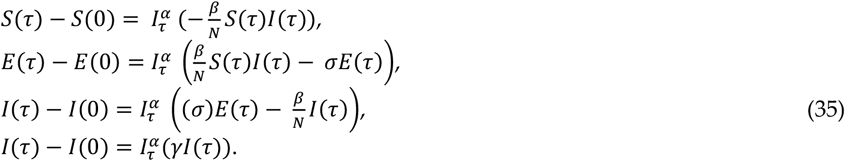

Using the condition (34), we obtained the system in terms of *F*_*i*_, *i* = 1,2,3,4 given as follows:

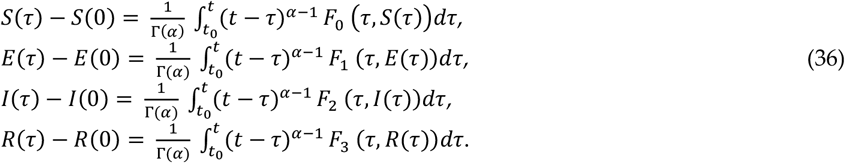

Using Picard iterations, equations (36) can be written as follows:

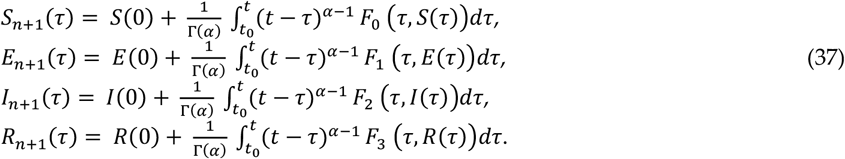

We consider *X*(*τ*) = (*S*(*τ*), *E*(*τ*), *I*(*τ*), *R*(*τ*))^*T*^ with *F* (*τ, X*(*τ*)) = (*F*_0_ (*τ, S*(*τ*)), *F*_1_ (*τ, S*(*τ*)), *F*_2_ (*τ, S*(*τ*)), *F*_3_ (*τ, S*(*τ*)))^*T*^ and 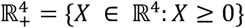.

#### Lemma 1

*If there is a solution to the system (31) with a positive initial condition, it will continue to be positive*.

**Proof**. In order to prove this result, we consider the initial condition (32) and we observe the following assertions hold:

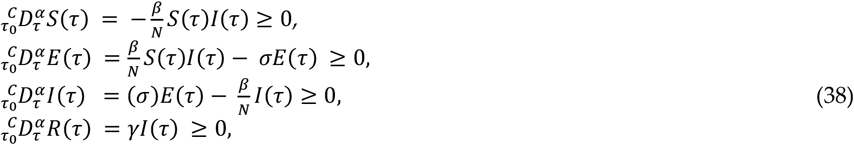

for all *τ* ∈ [0, *T*]. Using the statement mentioned in [24], we can further conclude that the solution *X*(*τ*) = (*S*(*τ*), *E*(*τ*), *I*(*τ*), *R*(*τ*))^*T*^ of the system (31) belongs to ℝ^4^, hence completes the proof. *□*

#### Lemma 2

*The function F*(*τ, X*(*τ*)) *defined in system (34) satisfied the Lipschitz condition, i,e*.; ||*F*(*τ, X*(*τ*) − *F*(*τ, X*^*\**^(*τ*)|| ≤ *β*|| (*X*(*τ*) − *X*^*\**^(*τ*))||, *where η* = max{|*β*|, |*β* + *σ*|, |σ+ *β*|, |(γ)|}.

**Proof**. Considering the couple functions *S*(*τ*) and *S*^*\**^(*τ*) such that the equality is given as follows:

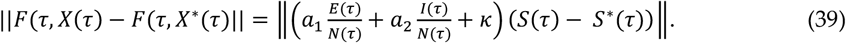

Suppose

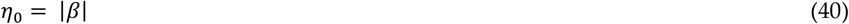

we deduce the following inequality:

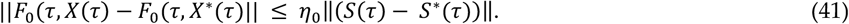

Similarly, we get the following:

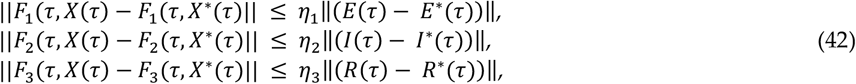

where

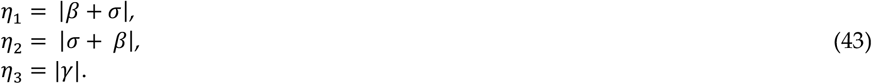

From equations (41-42), we show that all the kernels, *F*_0_, *F*_1_, *F*_2_, and *F*_3_, satisfy the Lipschitz condition. *□*

#### Theorem 1

*Using Lemma 2, the system (34) has a unique positive solution when*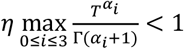.

**Proof**. We can write the solution of the system (34) as follows:

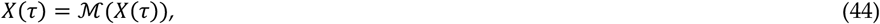

where *ℳ*: P([0, T], ℝ^4^) **→** P([0, T], ℝ^4^) is the Picard operator, hence follows

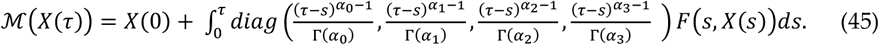

Then we have the following inequalities:

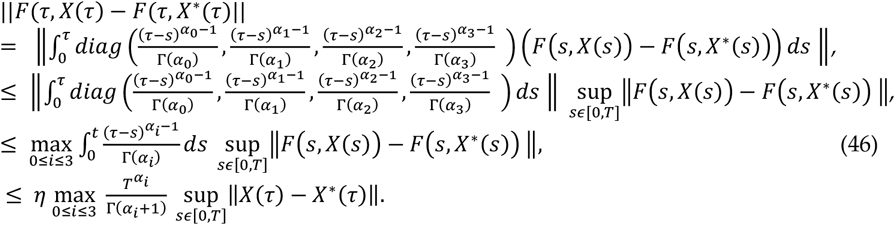

Since 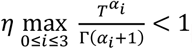, for *τ* ≤ *T*, and the operator *P* is a contraction. Hence, we prove that the system (34) has a unique solution.

## 5.Results

The basic reproduction number *R*_0_ of the model equation (31) is an essential parameter to acknowledge the basic dynamic behavior of the epidemic. It is the average number of cases generated by infectious individuals in the community in the absence of preventive measures [26]. In comparison to *R*_0_ the effective reproduction number *R*_*t*_ is used to calculate the number of individuals in a population that is entirely susceptible to infection and when control measures have been implemented. The effective reproduction number, which is calculated as an indicator of the virus’s transmissibility and represents the average number, is as follows:

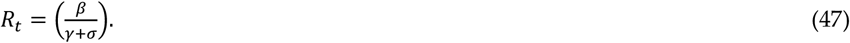

when *R*_*t*_ > 1, infection of the virus is regarded as spreading throughout the population as a whole. And *R*_*t*_ < 1 denotes a reduction in virus infection as a result of vaccination and other preventive measures. Using the parameters’ values from Table 1, we obtain *R*_*t*_ *≈* 0.099 < 1, indicating that the pandemic will disappear.

Using data from the Saudi Arabian Ministry of Health, we could use the curvature of the curve (the number of newly infected cases) to predict the dynamic behavior of the COVID19 pandemic in Saudi Arabia from 2020 to 2022 and from the start of 2023. The daily infection data are particularly unreliable since they rely so heavily on the changing test results that are made accessible every day. In this regard, the viral shading data in the wastewater may help to ensure the accuracy of the findings of individuals infected with the coronavirus. The second wave of COVID-19 infections began to spread in Saudi Arabia in December 2021, while the third wave came in January 2022. In the middle of December 2020, the first vaccine was given approval for usage in Saudi Arabia. The curve of daily COVID-19 infection cases in Saudi Arabia during the second wave (December 2021 to December 2022) of the outbreak is shown in Figure 4.

**Figure 4.**
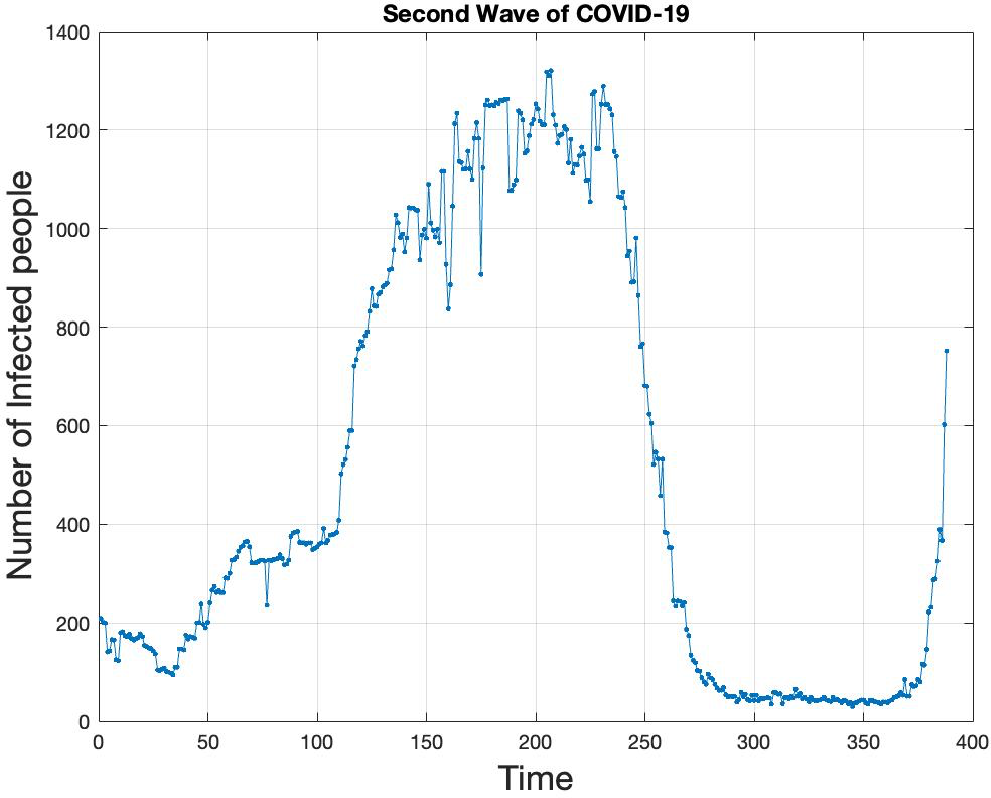
Number of COVID-19 infected cases during the second wave in Saudi Arabia.

Figure 5 (a) displays the simulation of the discrete time SEIR model (26) over the probability of viral load in wastewater for time *t* = 300 days from the start of the pandemic in Saudi Arabia.

**Figure 5.**
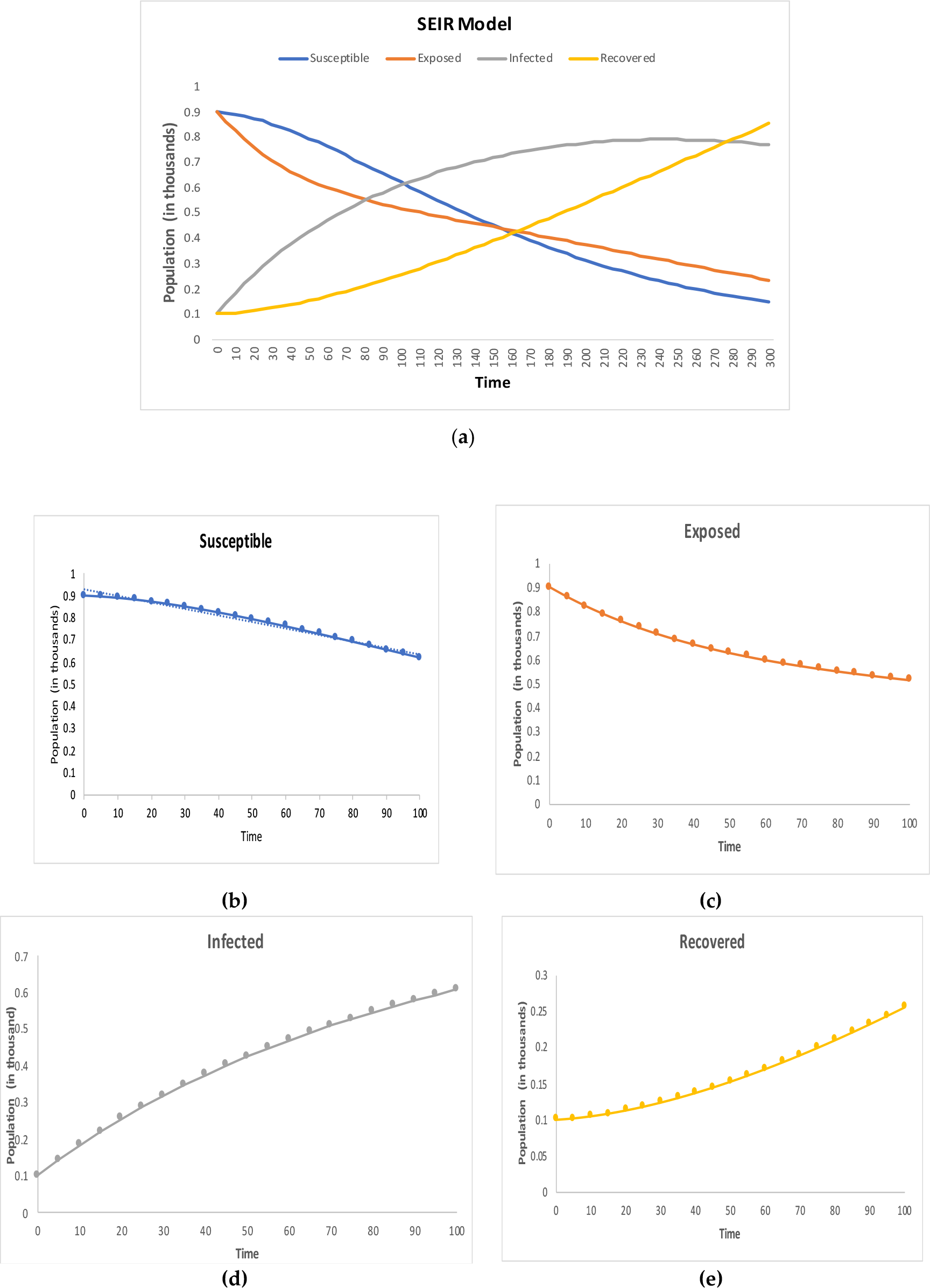
Simulation of (**a**) discrete time SEIR model (23) over time *t* = 300 ; (**b**) Susceptible over *t* = 100; (**c**) Exposed over *t* = 100, (**d**) Infected over *t* = 100, and (**e**) Recovered over *t* = 100 in case of *R*_*t*_ < 1.

Figure 5(b), (c), (d), and (e) show the simulation of the viral load in wastewater under susceptible, exposed, infected, and recovered respectively through time *t* = 100 days. Using MATLAB and the parameter values from Table 1, the fractional SEIR model (31) is simulated in case of *R*_*t*_ < 1 for various values of the fraction order. Figures 6, 7, 8, and 9 represent the dynamic behavior of fractional *S*(*t*), *E*(*t*), *I*(*t*), and *R*(*t*), respectively with different fractional values over time *t* = 100 days.

**Figure 6.**
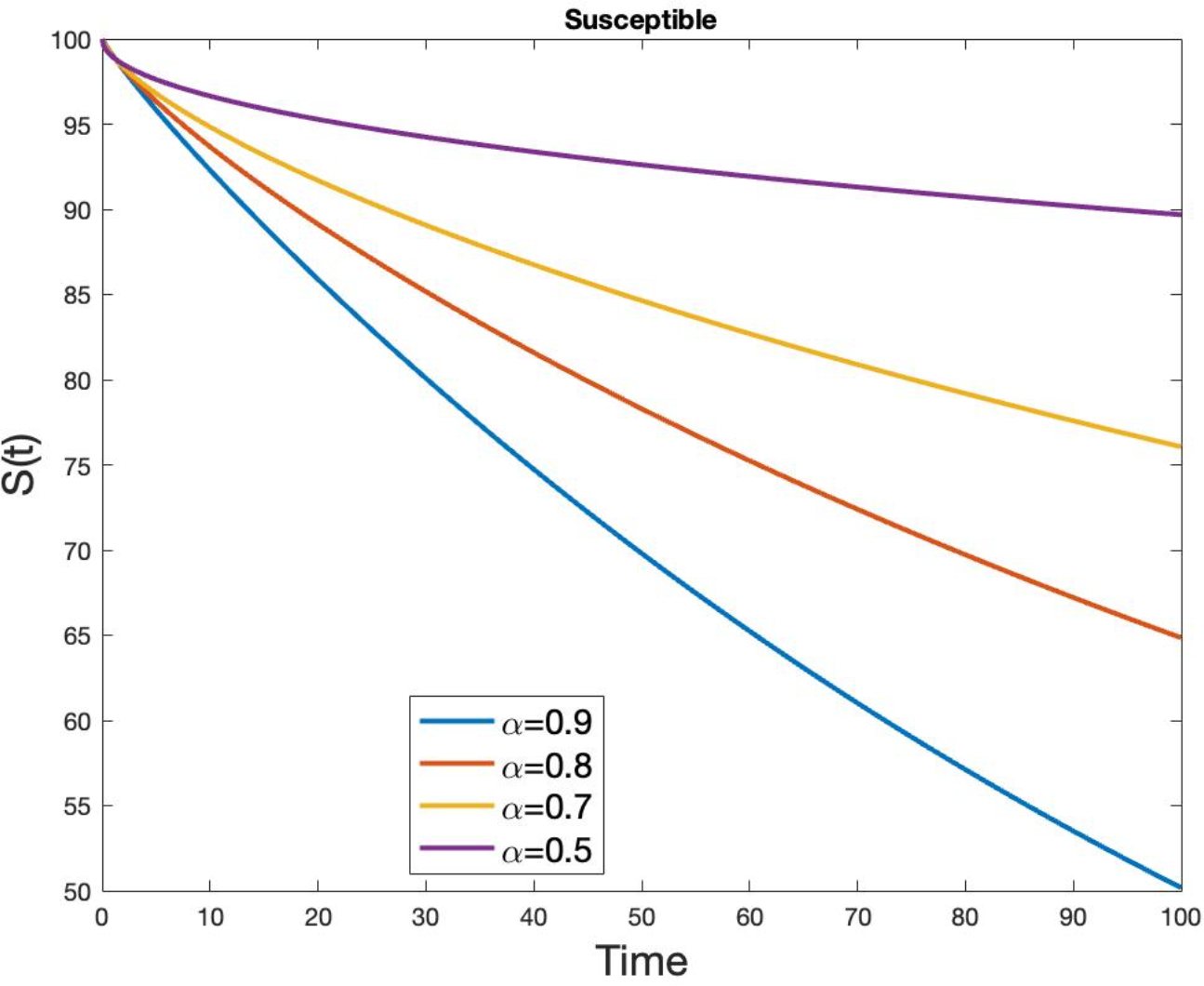
Simulation results for the dynamic behavior of susceptible *S*(*t*) at fractional order α = 0.9, α = 0.8, α = 0.7, and α = 0.5, in case of *R*_*t*_ < 1.

**Figure 7.**
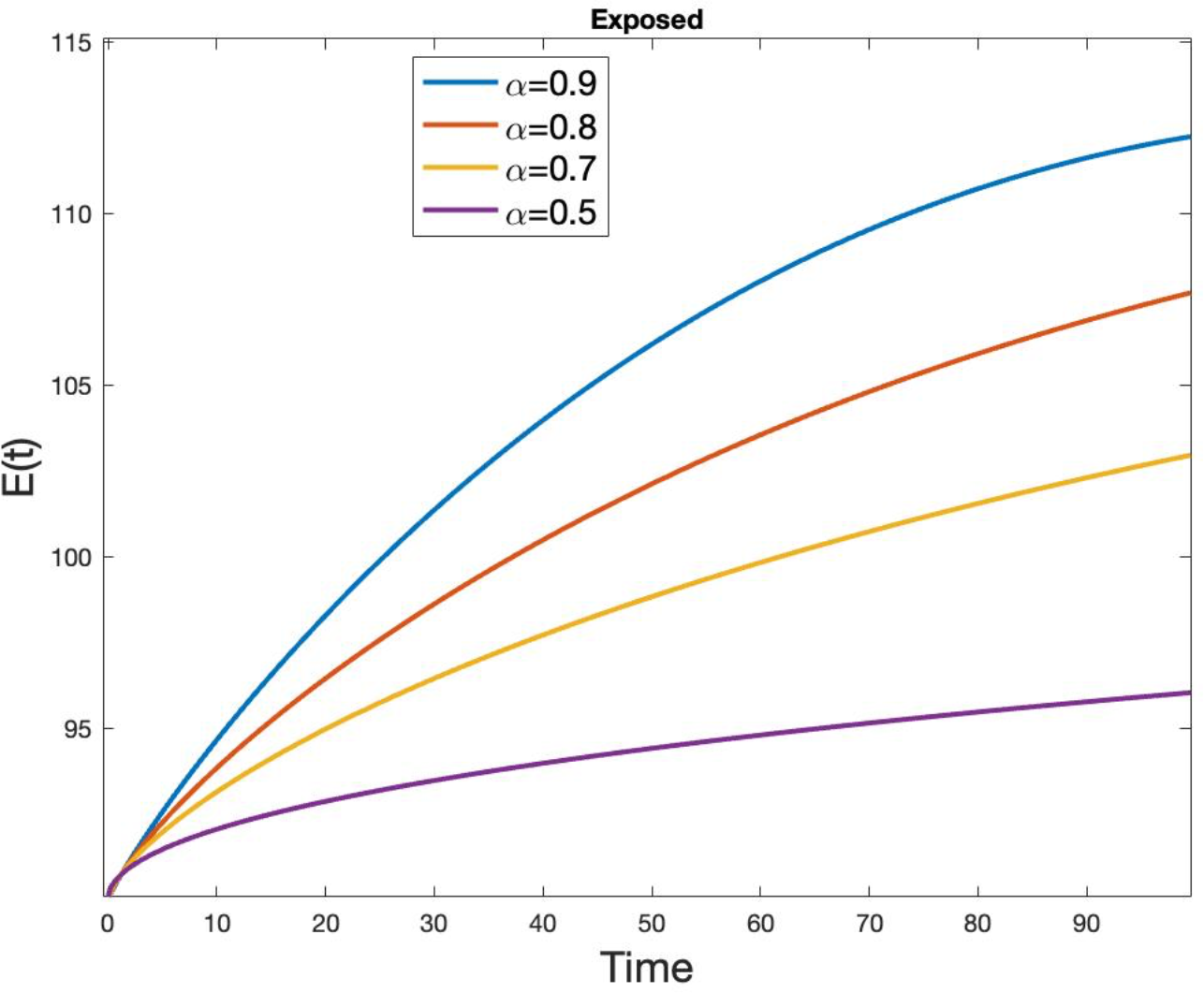
Simulation results for the dynamic behavior of Exposed *E*(*t*) at fractional order α = 0.9, α = 0.8, α = 0.7, and α = 0.5, in case of *R*_*t*_ < 1.

**Figure 8.**
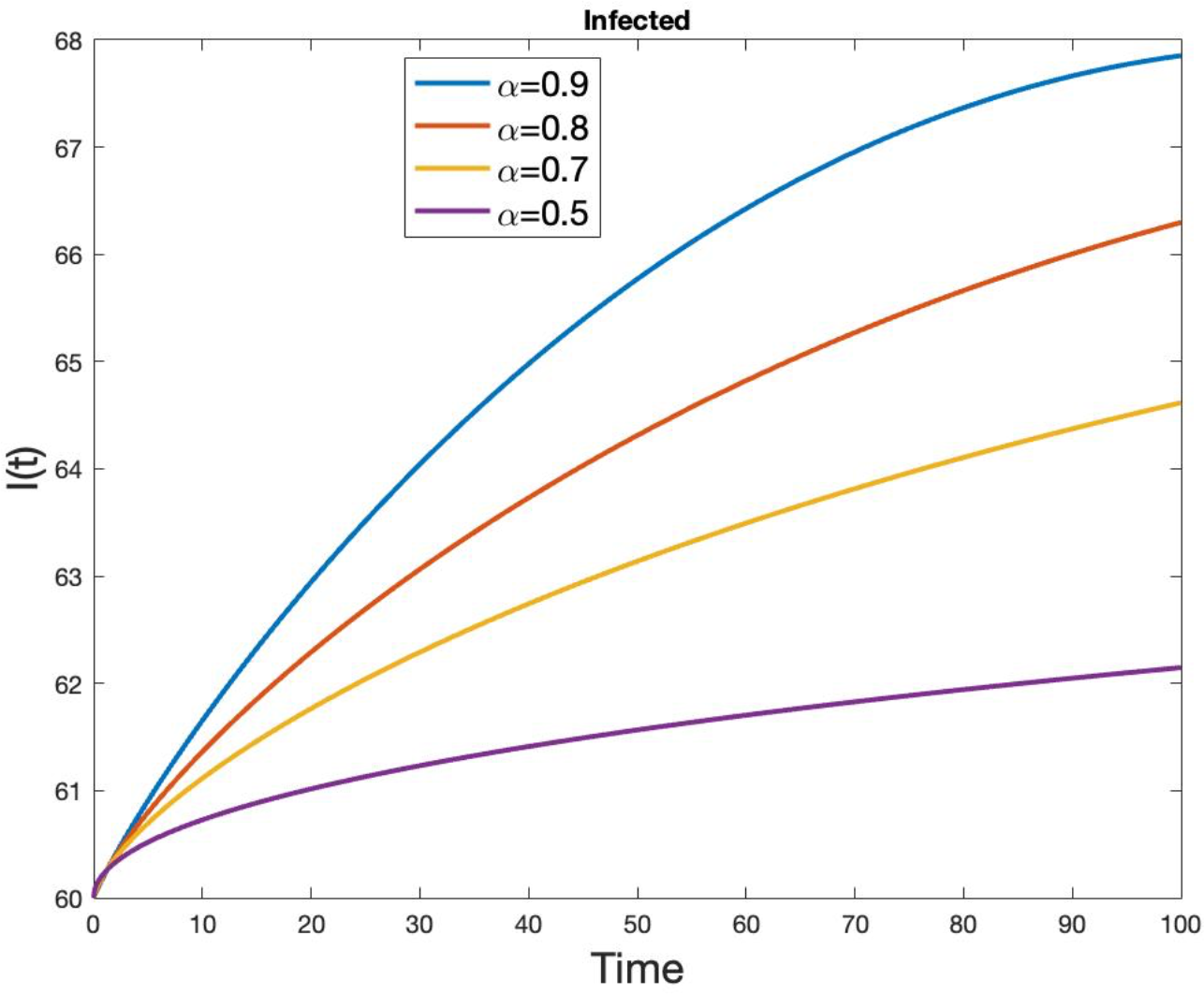
Simulation results for the dynamic behavior of Infected *I*(*t*) at fractional order α = 0.9, α = 0.8, α = 0.7, and α = 0.5, in case of *R*_*t*_ < 1.

**Figure 9.**
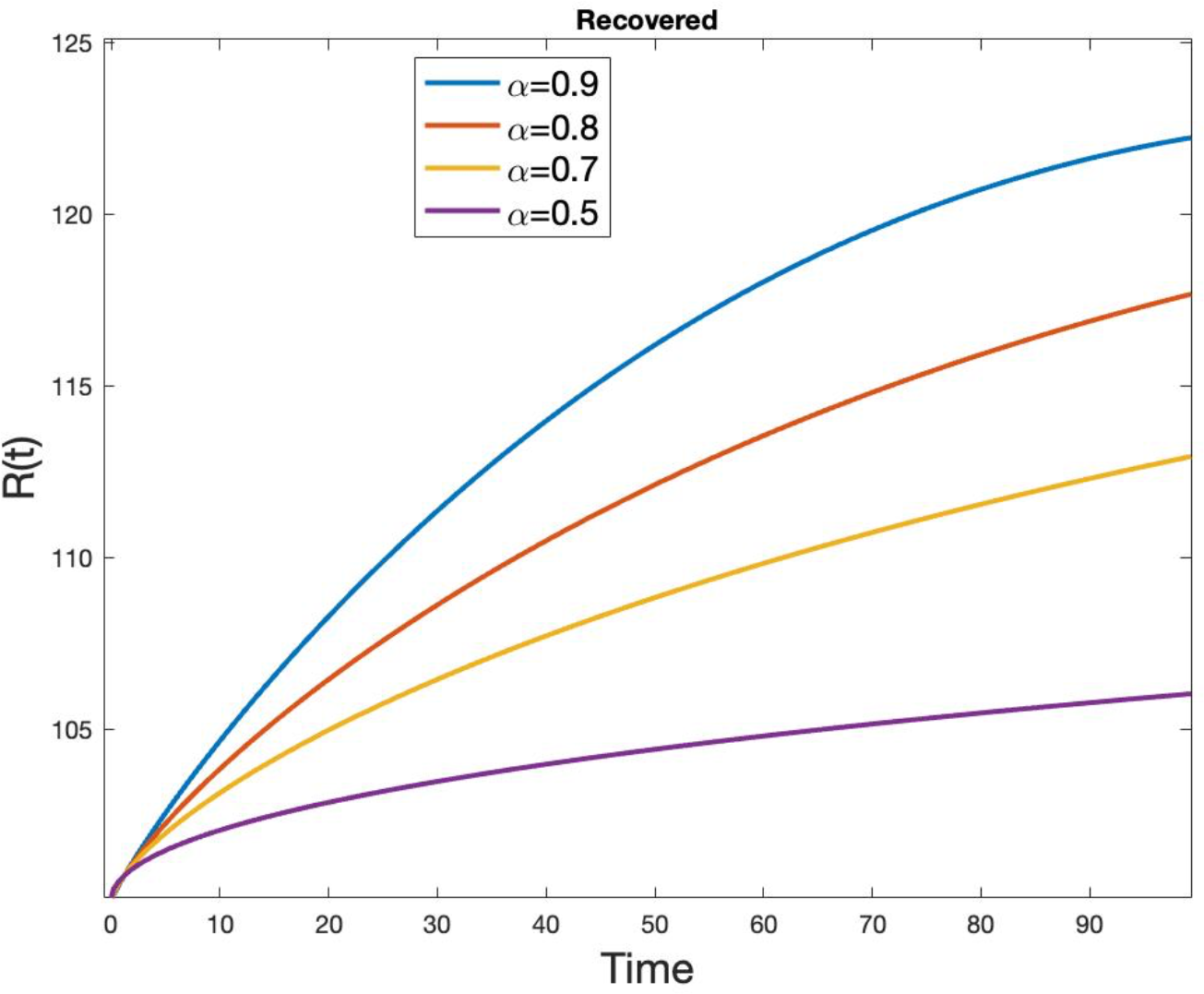
Simulation results for **t**he dynamic behavior of Recover *R*(*t*) at fractional order α = 0.9, α = 0.8, α = 0.7, and α = 0.9, in case of *R*_*t*_ < 1.

Furthermore, we simulate the fractional system (31) in the event that *R*_t_ > 1 because it is widely known that the epidemic will gradually spread in the population that is susceptible. We are able to change the curvature of the curve by altering the values of the fractional orders, as shown by the simulated figure with various fractional order values. Using the fractional ordered fractional order values of the SEIR model, we can independently alter the curves’ curvature. This fractional model’s proper gives us the ability to adjust the model to be better aligned with real data than integer models do.

The fractional SEIR model (31) simulation is performed for the effective reproduction number *R*_+_ > 1. Figures 10 (a), (b), (c), and (d) represent the dynamic behavior of fractional *S*(*t*), *E*(*t*), *I*(*t*), and *R*(*t*), respectively with different fractional values over time *t* = 100 days when the epidemic is gradually increasing. From Figure 10 above, we can see that by adjusting the fractional order, the simulation of the fractional-order SEIR model can better replicate the real-world process.

**Figure 10.**
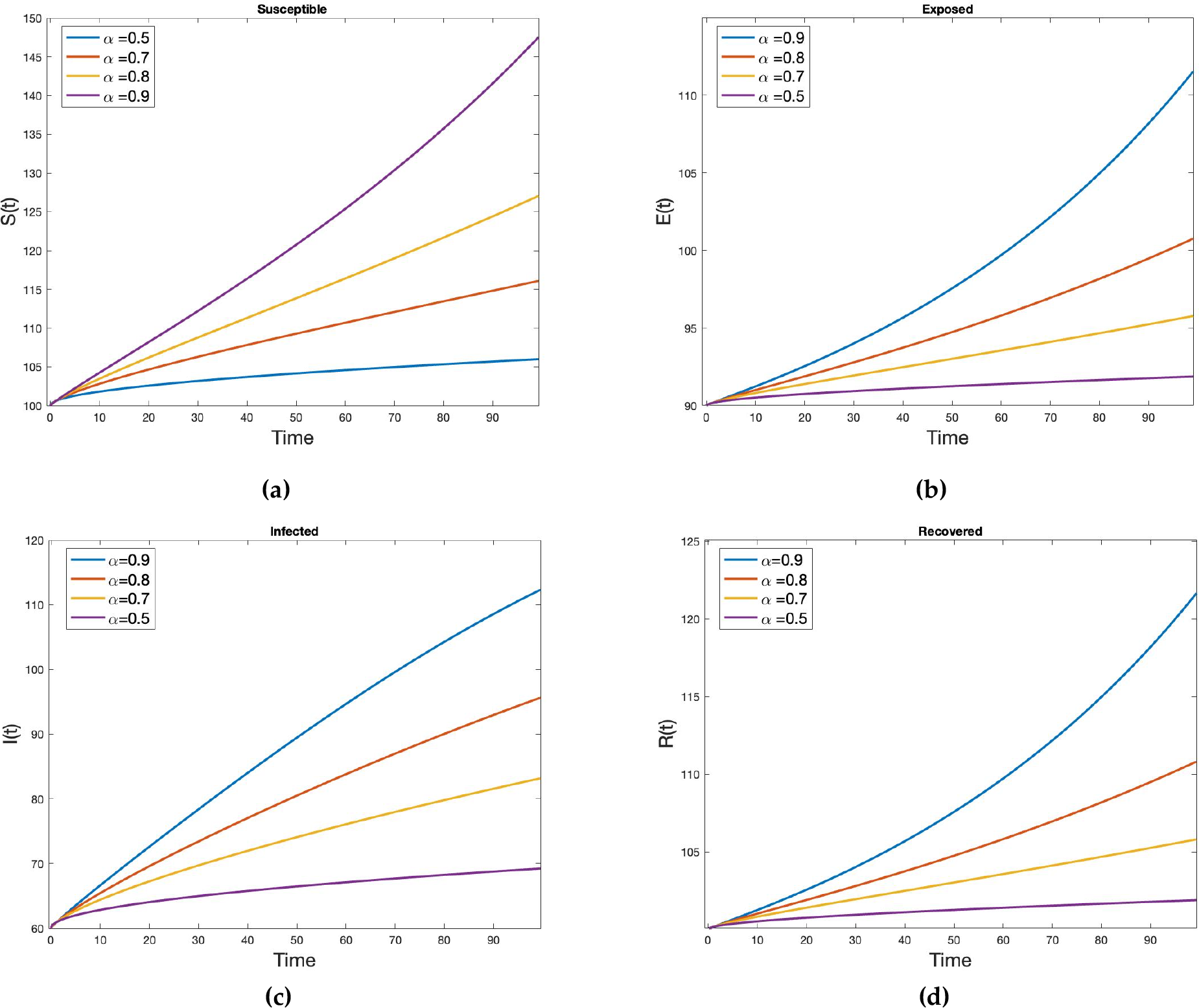
Simulation results for the dynamic behavior of (**a**) Susceptible *S*(*t*), (**b**) Exposed *E*(*t*), (**c**) Infected *I*(*t*), and (**d**) Recovered *R*(*t*) at fractional order α = 0.5, α = 0.7, α = 0.8, and α = 0.9, in case of *R*_*t*_ > 1.

The fractional-order model is useful for predicting the dynamics of an epidemic over a short period of time. We consider the statistics data for COVID-19 in Saudi Arabia from September 1, 2021, to December 30, 2021, which falls under the second wave. When taking into account the mean and standard deviation of the viral load in the untreated wastewater at wastewater treatment plants in Saudi Arabia, the fractional SEIR model is compared with the real data.

For comparing the simulation of the fractional SEIR model, the daily infected and recovered cases during the second coronavirus wave in Saudi Arabia are taken into consideration. Figure 11 displays the simulation results at various fractional orders for both infection and recovery cases. We can observe how the dynamic properties of the SEIR model are influenced by the fractional ordering of the derivatives. The fractional order α variation can offer a better acceptable model that is consistent with the actual data for estimating the number of cases that will be infected. The adjustment in fractional order also produces findings that are more consistent with the actual data when forecasting recovery situations.

**Figure 11.**
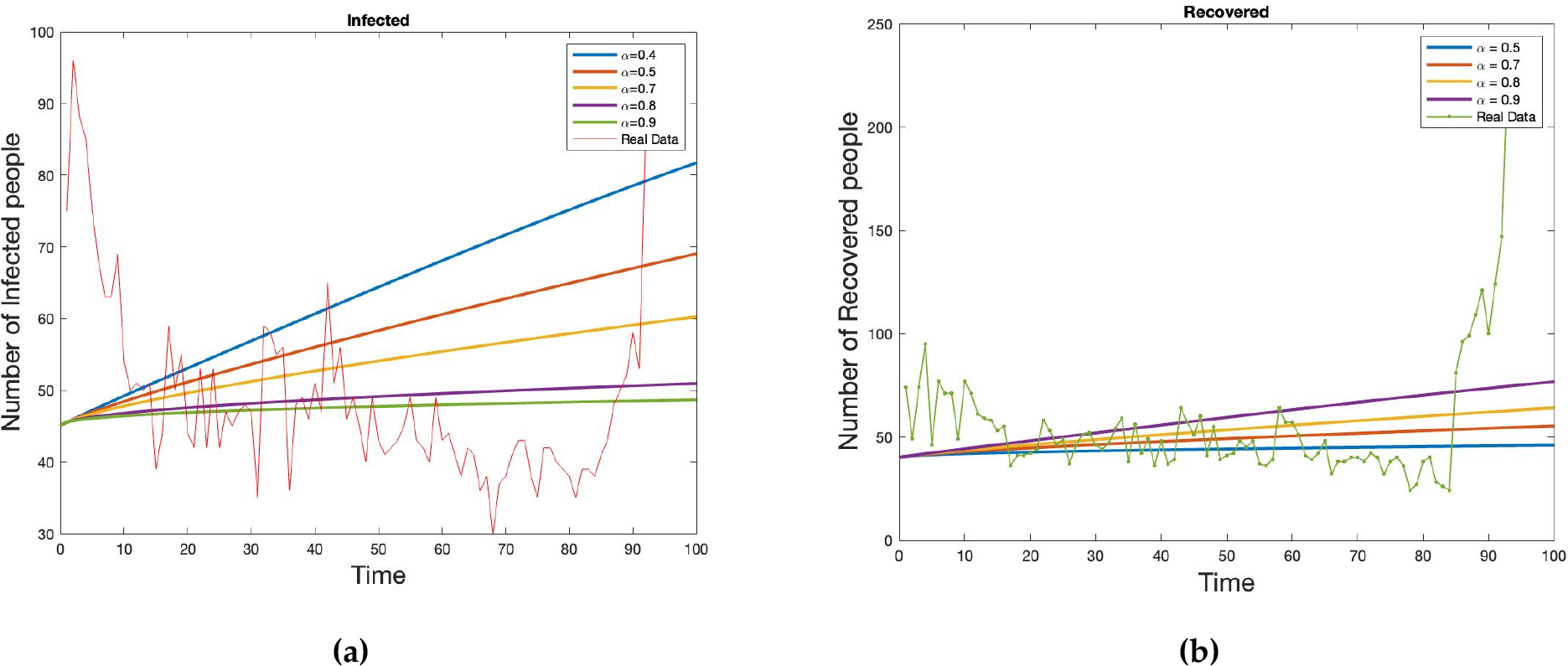
Comparison of the fractional SEIR model simulation at various fractional order for (**a**) *I*(*t*), in relation to the infected cases, and (**b**) *R*(*t*), in relation to the recovered cases from September 1, 2021 to December 30, 2021.

## 6. Conclusions

In this study, we examine the dynamic behavior of COVID-19 infection using possible information gathered from wastewater treatment facilities in Saudi Arabia’s major cities. Since SARS-CoV-2 spreads not only through the air but also through sewage water, it is essential to study the untreated wastewater that reaches wastewater treatment facilities. To fully understand the potential coronavirus load in treatment wastewater and its spread in the community, we apply mathematical modeling, such as the log-normal model and fractional SEIR model. The log-normal model estimates the potential viral load in wastewater, which helps determine the time of the pandemic’s potential maximum outbreak. The existence and uniqueness of positive solutions of the fractional SEIR model are derived.

It is challenging to get the integer model to accept specific statistics (real data) when dealing with the model. This is due to the constant curvature, which prevents the integer model from agreeing with the gathered statistics of real data. These challenges are overcome through the use of fractional order models. By controlling the curvature of each state independently, a fractional order SEIR model has improved the system’s outcomes and made forecasts more accurate.

In conclusion, our model simulates the process of predicting COVID-19 infection through wastewater. Wastewater surveillance and clinical testing data should be merged in order to assess typical population-level viral shedding and possible future viral transmission with greater accuracy. Although wearing masks, getting vaccinated, and restricting mobility may help, viral shedding data from wastewater treatment plants may be one way to prevent viral propagation into the community and potential future discovery.

## Data Availability Statement

All the data used in this paper are available on the website of the Ministry of Health, Saudi Arabia (https://covid19.moh.gov.sa/, accessed on 11 July 2023), and on the World Health Organization (WHO) website (https://covid19.who.int/data, accessed on 11 July 2023).

## Acknowledgments

The authors extend their appreciation to the Deputyship for Research and Innovation, Ministry of Education in Saudi Arabia, for funding this research work through project number ISP22-6.

